# A novel multimodal approach to improve upper extremity function after moderate-to-severe stroke

**DOI:** 10.1101/2023.04.23.23288971

**Authors:** Amit Sethi, Alvaro Pascual-leone, Emiliano Santarnecchi, Ghaleb Almalki, Chandramouli Krishnan

## Abstract

**Background:** Interventions to recover upper extremity (UE) function after moderate-to-severe stroke are limited. Transcranial random noise stimulation (tRNS) is an emerging non-invasive technique to improve neuronal plasticity and may potentially augment functional outcomes when combined with existing interventions, such as functional electrical stimulation (FES).

**Objective:** The objective of this study was to investigate the feasibility and preliminary efficacy of combined tRNS and FES-facilitated task practice to improve UE impairment and function after moderate-to-severe stroke.

**Methods:** Fourteen individuals with UE weakness were randomized into one of two groups: 1) tRNS with FES-facilitated task practice, or 2) sham-tRNS with FES-facilitated task practice. Both groups involved 18 intervention sessions (3 per week for 6 weeks). We evaluated number of sessions completed, adverse effects, participant satisfaction, and intervention fidelity between two therapists. UE impairment (Fugl-Meyer Upper Extremity, FMUE), function (Wolf Motor Function Test, WMFT), participation (Stroke Impact Scale hand score, SIS-H), and grip strength were assessed at baseline, within 1 week and 3 months after completing the intervention.

**Results:** All participants completed the 18 intervention sessions. Participants reported minimal adverse effects (mild tingling in head). The two trained therapists demonstrated 93% adherence and 96% competency with the intervention protocol. FMUE and SIS-H improved significantly more in the tRNS group than in the sham-tRNS group at both timepoints (p≤0.05), and the differences observed exceeded the clinically meaningful differences for these scores. The WMFT and paretic hand grip strength improved in both groups after the intervention (p≤0.05), with no significant between group differences.

**Conclusion:** Our findings show for the first time that combining tRNS and FES-facilitated task practice is a feasible and promising approach to improve UE impairment and function after moderate-to-severe stroke.

## 1. Introduction

Over two-thirds of stroke survivors experience moderate-to-severe impairments in their paretic upper extremity (UE),(Tsao et al., 2022) making it difficult for them to perform daily tasks.(Crichton et al., 2016) Existing combinations of neuromodulatory interventions such as repetitive transcranial magnetic stimulation (r-TMS) or transcranial direct current stimulation (tDCS) with functional electrical stimulation (FES) or robot-assisted task practice have limited effectiveness in improving hand function in this population.(Monte-Silva et al., 2019; Reis et al., 2021; Sun et al., 2021) Transcranial random noise stimulation (tRNS) employs stochastic resonance to modulate motor cortical excitability.(van der Groen et al., 2022) Stochastic resonance amplifies weak brain activity by increasing signal-to-noise ratio and can promote longer-lasting changes in neuronal plasticity.(Arnao et al., 2019; Terney et al., 2008; van der Groen et al., 2022) This study aimed to examine the feasibility and preliminary efficacy of combining tRNS with FES-facilitated task practice on improving UE impairment and function after moderate-to-severe stroke.

## 2. Methods

### 2.1 Design

This was a single-center, parallel-group, double-blind pilot randomized controlled clinical trial. Participants were randomized using simple randomization by a coordinator to receive six weeks (3×/week) of: 1) tRNS with FES-facilitated task practice, or 2) sham-tRNS with FES-facilitated task practice. Demographics provided in Supplementary Table 1. A licensed occupational therapist, blinded to the intervention protocol, evaluated the participants before and after the intervention. Participants were blinded to the intervention using an established sham-tRNS protocol.(Terney et al., 2008)

### 2.2 Participants

Participants (n=14, male=9, female=5, age=63.2 ± 5 years) were included if they: 1) were ≥ 21 years; 2) had chronic (> 6 months) unilateral hemiparesis due to a stroke; and 3) < 20° degrees of active wrist extension indicating moderate-to-severe impairments. Exclusion criteria were severe aphasia, severe spasticity (Modified Ashworth Scale ≥ 3), contraindications of brain stimulation,(Rossi et al., 2021) and cognitive impairment (Mini-Mental Status Examination< 23). Participants provided written informed consent approved by the University of Pittsburgh Institutional Review Board.

### 2.3 Intervention

Of the 95 participants screened, 35 met the study criteria of which only 14 provided consent and completed the study as per the protocol. See Figure 1 for a summary including reasons for exclusion.

**Fig 1a.**
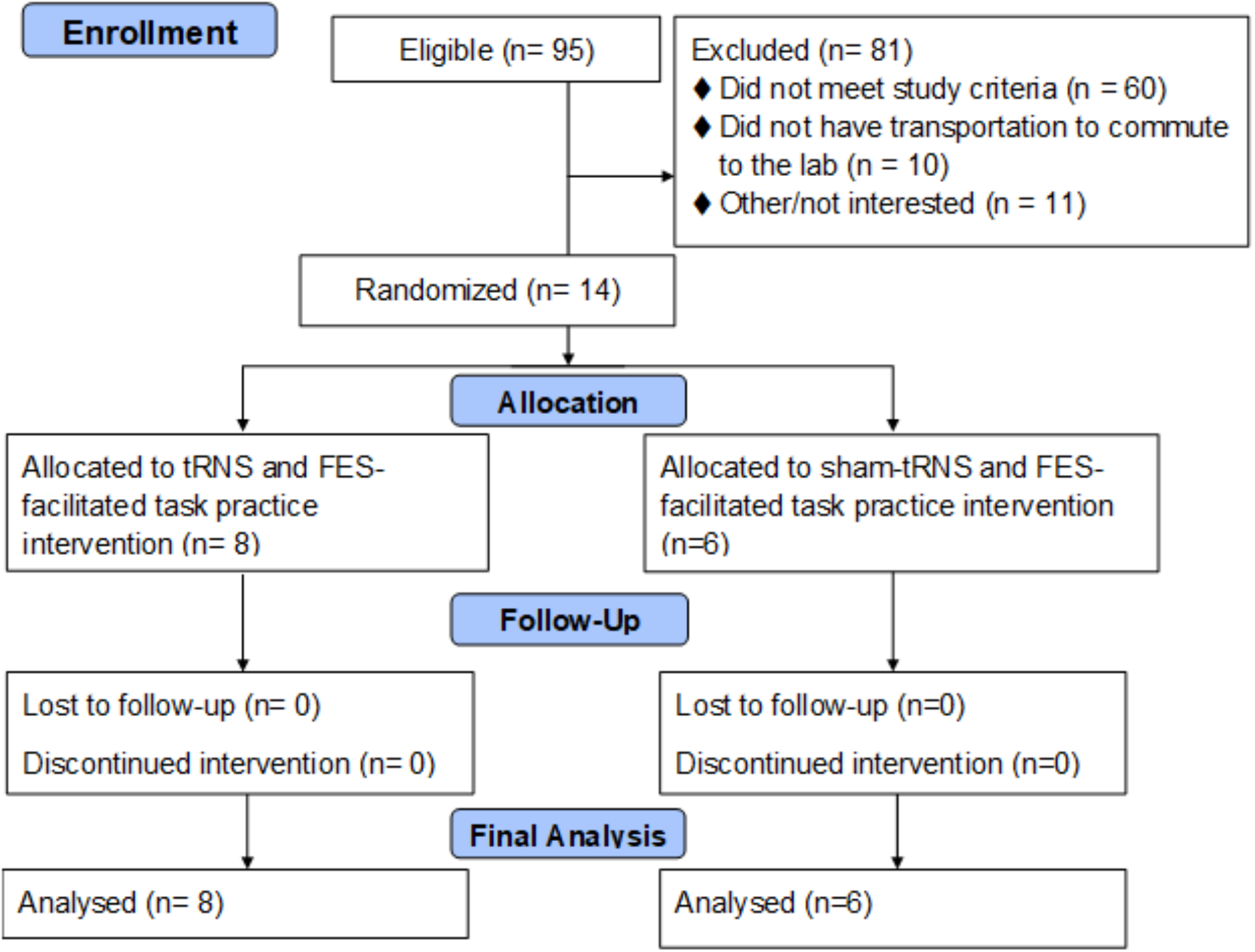
Study CONSORT (CONsolidated Standards of Reporting Trials) flow diagram, illustrating the screening, enrollment, and trial design of this clinical trial.

**Fig 1b.**
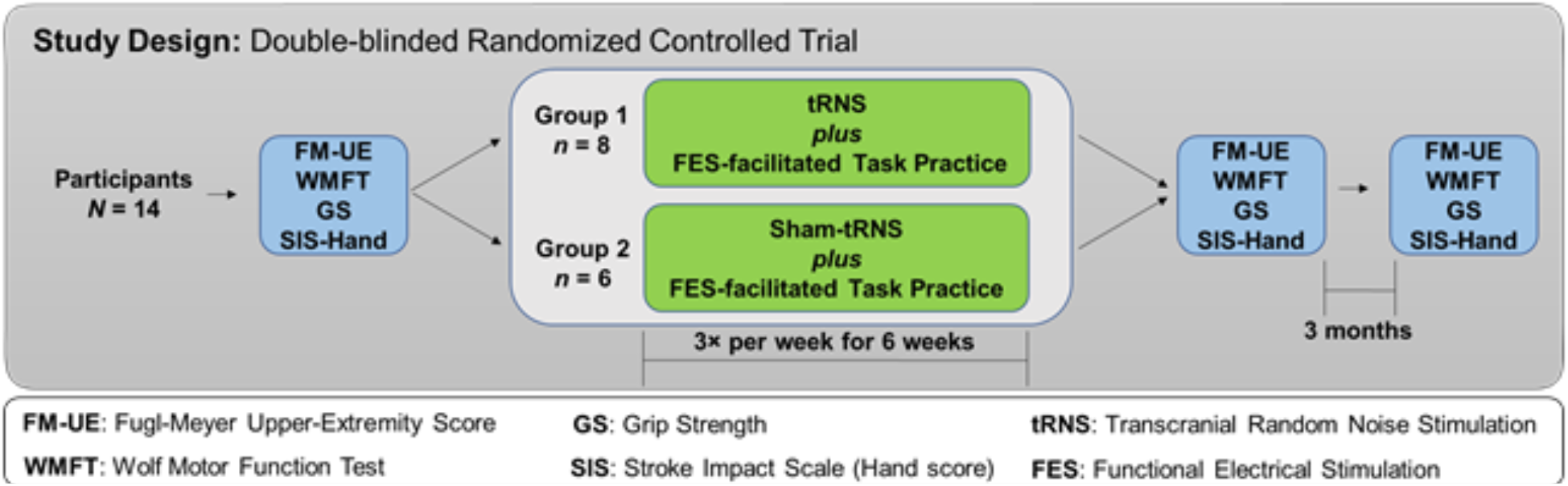
A schematic of the study protocol illustrating the testing and the training visits associated with this study.

#### Trns

tRNS (2mA, 100-500Hz) was applied using the StarStim device (Neuroelectrics Barcelona SL) via four NG pi-stim electrodes (1cm radius). The active electrodes were placed over the ipsilesional motor cortex (C3 or C4) and the return electrodes were placed over the contralesional hemisphere (PO8, CP6, and F4 or PO7, CP5, and F3) such that the resulting electric field had a maximum induced current in the hand knob of the central sulcus (Figure 2a).

**Figure 2:**
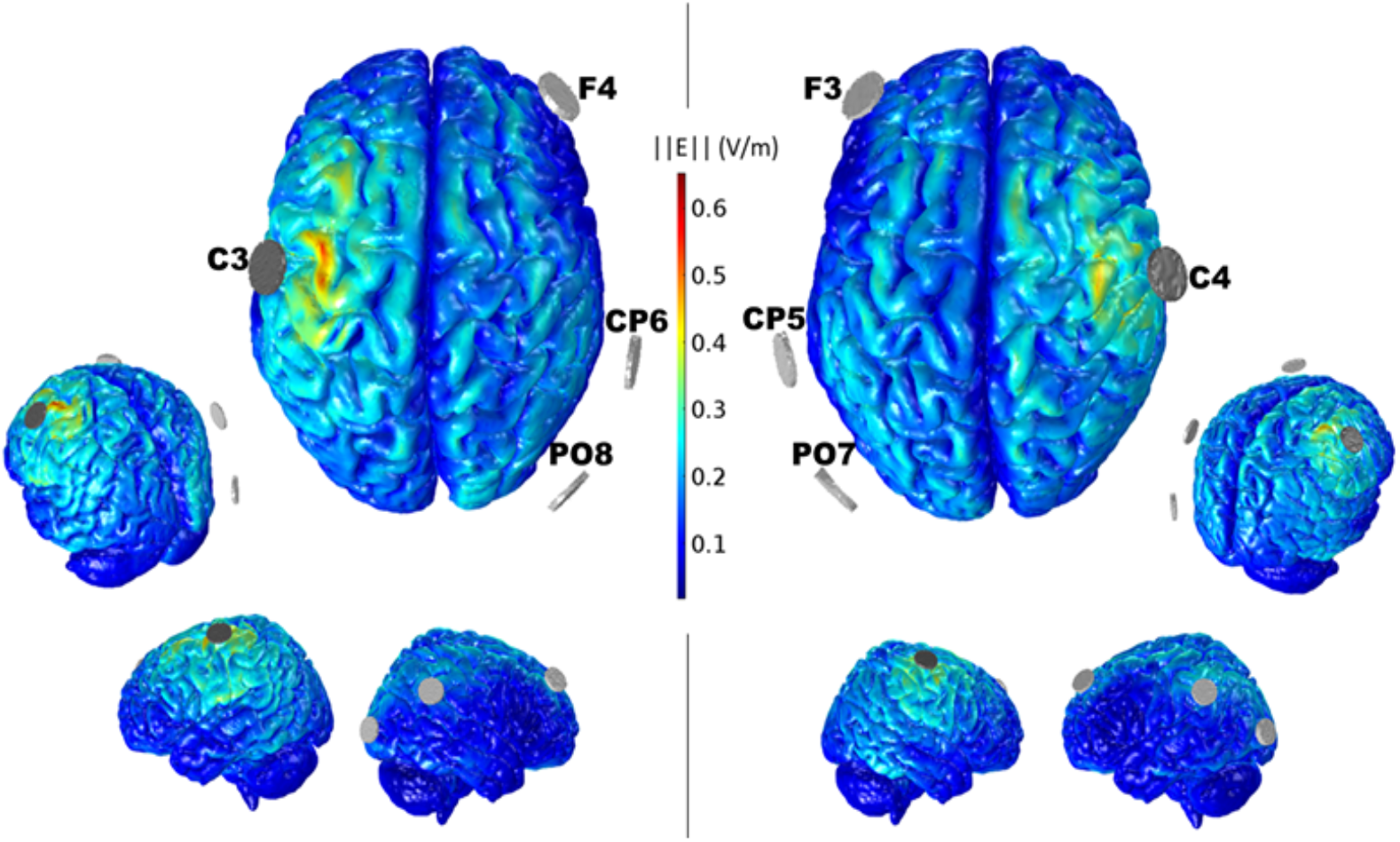
The general electric field plots generated by tRNS show the electric field norm distribution (V/m) on a template cortical surface induced by the two montages used in this study. The main electrode (C3 or C4) outputs the peak value for the current (2mA), and each return electrode on the opposite hemisphere represents one third of this value. The electric field was calculated in Comsol (www.comsol.com), by solving Laplace’s equation for the electrostatic potential, on the template head model.

#### FES-facilitated task practice

A U.S. FDA-approved FES device (Neuromove, Zynex, Denver, CO) was used to deliver task practice using an established protocol.(Monte-Silva et al., 2019) The FES delivered stimulation between 15 to 29 mA using a biphasic waveform for 5 seconds at 50 Hz. The intervention started with a 15-minute block of stimulating hand flexors to facilitate grasping objects, followed by a second 15-minute block of stimulating hand extensors to facilitate releasing objects. These two blocks were repeated for the last half hour of the intervention. Participants performed 150-160 repetitions per session.

#### Behavioral strategies

A behavioral contract was executed to emphasize paretic UE use at home. Participants kept weekly UE-use diary, rated paretic UE use on the Motor Activity Log,(Uswatte et al., 2005) discussed barriers, and reviewed weekly changes in performance. A videotape of the intervention sessions was shown every week to boost confidence in hand usage.

### 2.4 Outcome measures

Outcomes were evaluated at two timepoints (immediately [POST] and 3 months [POST-2] after the intervention). The primary outcome measure was Fugl-Meyer Upper Extremity Scale (FMUE), which measures the change in UE impairment. The secondary outcome measures included Wolf Motor Function Test (WMFT), grip strength, and the Stroke Impact Scale hand score (SIS-H).

### 2.5 Statistical Analysis

The effect of tRNS on UE function was evaluated using a Linear mixed model with group (tRNS, sham-tRNS), time (PRE, POST, POST-2), and group × time as fixed factors and subjects as a random factor. The baseline scores in each of the outcome measures were used as a fixed covariate in the model. Relevant post-hoc analyses with Šidák corrections were conducted. The alpha level was set to p*≤*0.05.

## 3. Results

All 14 enrolled participants completed the study as per the protocol, and no participant reported any adverse effects other than the common mild adverse effects of tRNS(van der Groen et al., 2022) (i.e., mild tingling) and FES-facilitated task practice (i.e., fatigue in the paretic UE). Participants also rated their satisfaction as high. The therapists demonstrated 93% adherence and 96% competency with the intervention protocol.

### Fugl-Meyer Upper Extremity Scale (FMUE, primary outcome measure)

A significant group × time interaction effect was observed for the upper-extremity FMUE (F = 3.348, p = 0.05, Figure 3). Posthoc analyses indicated that the FMUE was significantly higher in the tRNS group than in the sham-tRNS group at both time points (p = 0.046 and p = 0.003; Figure 1). Posthoc analyses also indicated that the tRNS group exhibited a significant increase from baseline in the FMUE at both POST (Δ = 6.75 ± 1.56, p < 0.001) and POST-2 (Δ = 8.75 ± 1.41, p < 0.001) timepoints, whereas the sham-tRNS group exhibited no significant improvements in the FMUE at both POST (Δ = 3.17 ± 1.80, p = 0.240) and POST-2 timepoint (Δ = 3.17 ± 1.63, p = 0.179).

**Figure 3:**
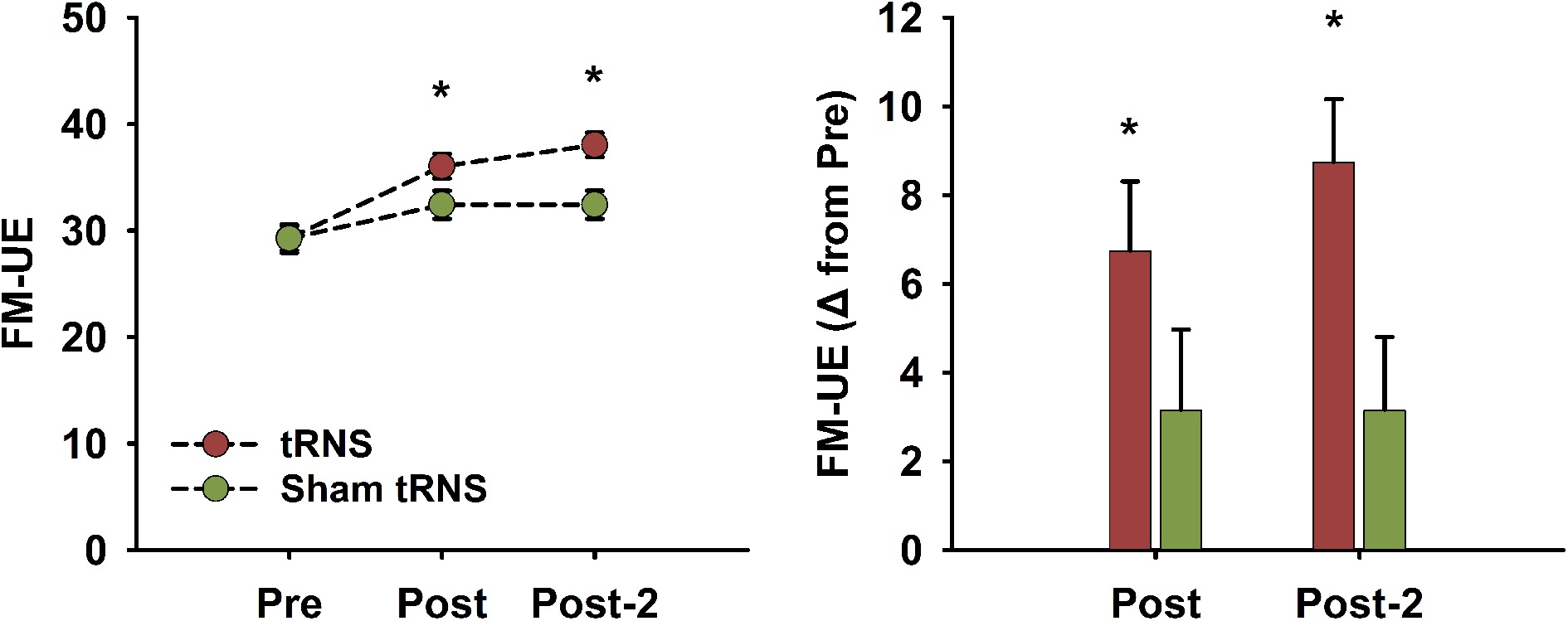
(i) Line plots showing the adjusted means of Fugl Meyer Upper Extremity Scale before (Pre), 1 week (Post), and 3 months (Post2) after the intervention in the tRNS plus FES-facilitated task practice and sham-tRNS plus FES-facilitated task practice and sham-tRNS group. (ii) Bar graphs showing the mean changes in Fugl Meyer Upper Extremity Scale from baseline in the tRNS plus FES-facilitated task practice and sham-tRNS plus FES-facilitated task practice and sham-tRNS group. Asterisks indicate significant differences at p</= 0.05 and error bars depict standard error of the mean.

### Wolf Motor Function Test (WMFT, secondary outcome measure)

A significant time effect was observed for the WMFT (F = 5.408, p = 0.011, Figure 4). Posthoc analyses indicated that the WMFT significantly improved at POST-2 (Δ = − 9.82 ± 3.0 seconds, p = 0.009) from baseline but not at POST (Δ = − 7.5 ± 3.5 seconds, p = 0.116). There were no significant group (F = 0.001, p = 0.974) or group × time interaction effect (F = 0.605, p = 0.554). *Grip Strength (secondary outcome measure)*

**Figure 4:**
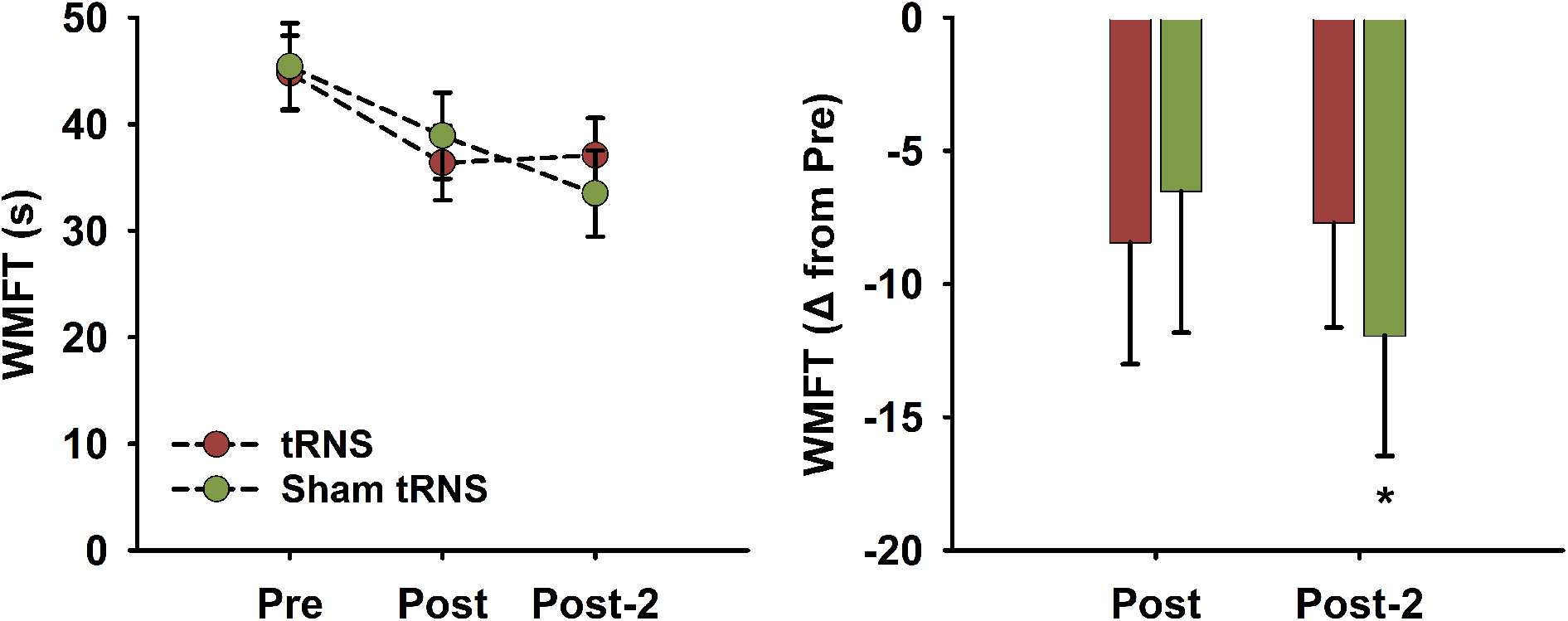
(i) Line plots showing the adjusted means of Wolf Motor Function Test before (Pre), 1 week (Post), and 3 months (Post2) after the intervention in the tRNS plus FES-facilitated task practice and sham-tRNS plus FES-facilitated task practice and sham-tRNS group. (ii) Bar graphs showing the mean changes in Wolf Motor Function Test from baseline in the tRNS plus FES-facilitated task practice and sham-tRNS plus FES-facilitated task practice and sham-tRNS group. Asterisks indicate significant differences at p</= 0.05 and error bars depict standard error of the mean.

A significant time effect was observed for the paretic hand grip strength (F = 5.188, p = 0.011, Figure 5). Posthoc analyses indicated that the paretic hand grip strength significantly improved at both POST (Δ = 3.53 ± 1.28 kg, p = 0.027) and POST-2 (Δ = 3.32 ± 1.13, p = 0.017) from baseline. There were no significant group (F = 0.523, p = 0.474) or group × time interaction effect (F = 0.395, p = 0.677).

**Figure 5:**
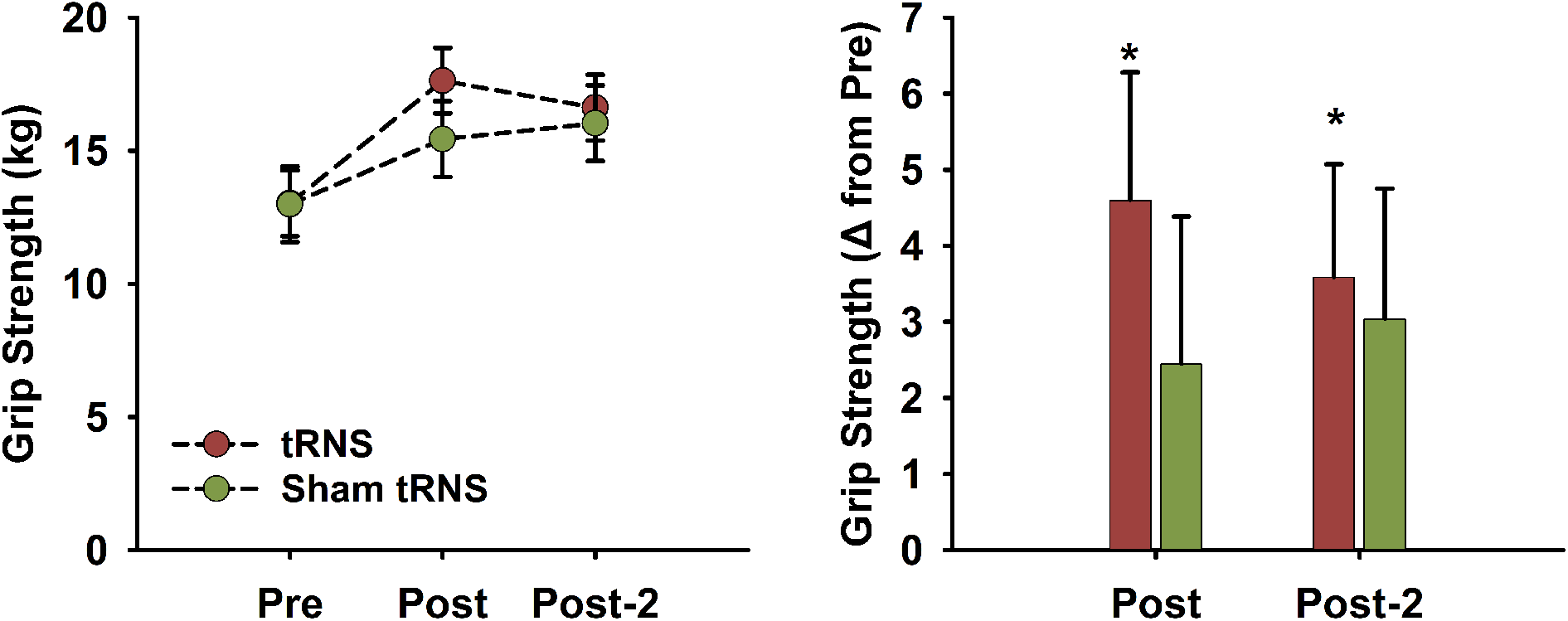
(i) Line plots showing the adjusted means of Grip Strength before (Pre), 1 week (Post), and 3 months (Post2) after the intervention in the tRNS plus FES-facilitated task practice and sham-tRNS plus FES-facilitated task practice and sham-tRNS group. (ii) Bar graphs showing the mean changes in Grip Strength from baseline in the tRNS plus FES-facilitated task practice and sham-tRNS plus FES-facilitated task practice and sham-tRNS group. Asterisks indicate significant differences at p</= 0.05 and error bars depict standard error of the mean.

### Stroke Impact Scale Hand Score (SIS-H, secondary outcome measure)

A significant group × time interaction effect was observed for the SIS-H (F = 3.583, p = 0.041). Posthoc analyses indicated that the SIS-H was significantly higher in the tRNS group than in the sham-tRNS group at both time points (p = 0.001 and p = 0.035; Figure 6). Posthoc analyses also indicated that the tRNS group exhibited a significant increase from baseline in the SIS-H at both POST (Δ = 6.13 ± 1.26, p < 0.001) and POST-2 (Δ = 4.63 ± 1.42, p = 0.011) timepoints, whereas the sham-tRNS group exhibited no significant improvements in the SIS-H at both POST (Δ = 1.00 ± 1.46, p = 0.873) and POST-2 timepoint (Δ = 1.5 ± 1.64, p = 0.749).

**Figure 6:**
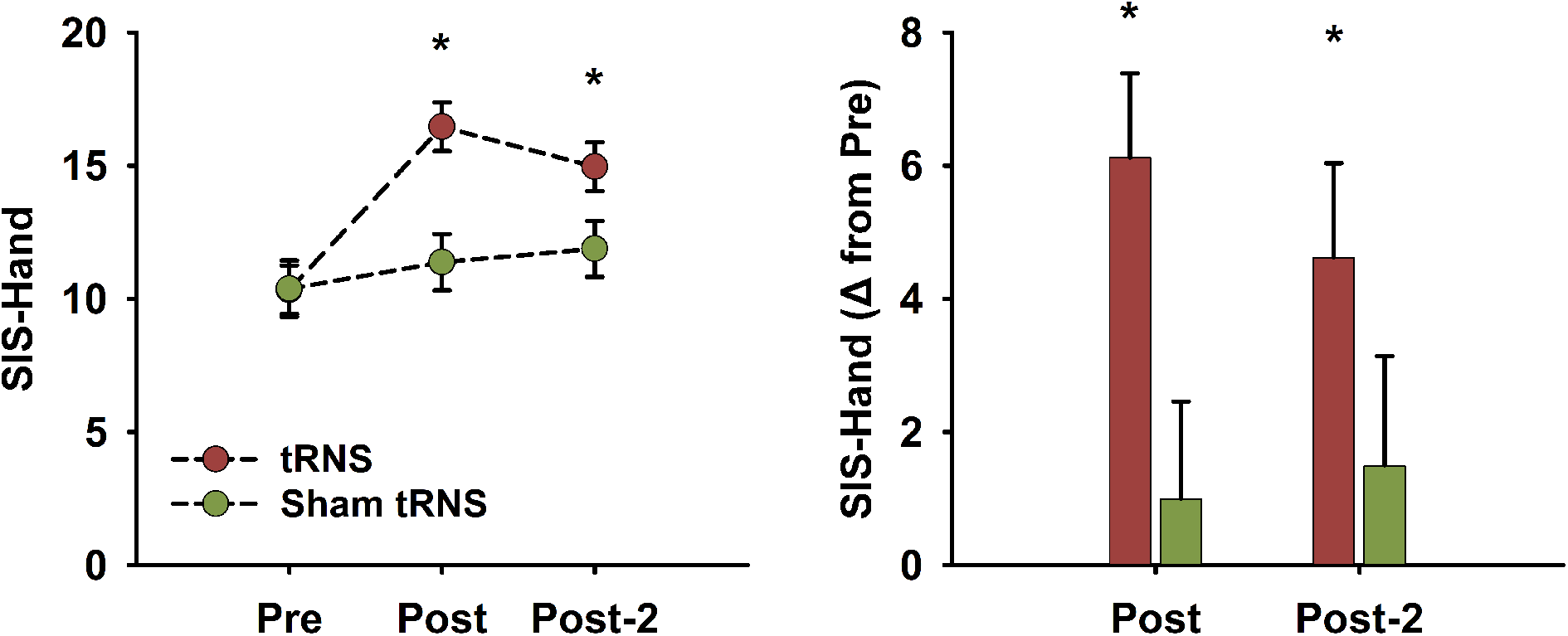
(i) Line plots showing the adjusted means of Stroke Impact Scale Hand score before (Pre), 1 week (Post), and 3 months (Post2) after the intervention in the tRNS plus FES-facilitated task practice and sham-tRNS plus FES-facilitated task practice and sham-tRNS group. (ii) Bar graphs showing the mean changes in Stroke Impact Scale Hand score from baseline in the tRNS plus FES-facilitated task practice and sham-tRNS plus FES-facilitated task practice and sham-tRNS group. Asterisks indicate significant differences at p</= 0.05 and error bars depict standard error of the mean.

## Discussion

This pilot randomized controlled study investigated the feasibility and sought to gather preliminary insights on the efficacy of combining tRNS and FES-facilitated task practice in improving UE impairment and function in individuals with moderate-to-severe chronic stroke. Results show outstanding tolerance and adherence, and significant improvements in UE impairment (FMUE) and hand function (SIS-H) at the end and after 3 months of intervention in the tRNS compared to sham tRNS and FES-facilitated task practice. These gains exceeded the minimal detectable, clinically-relevant change of these measures, and the combined intervention was safe. Our findings provide preliminary evidence for the first time showing that combined tRNS and FES-facilitated task practice can significantly improve UE impairment and hand function in individuals with moderate-to-severe stroke for whom currently there are no effective interventions. Future studies with larger sample size and mechanistic evaluations are needed to establish the effectiveness of this intervention.

## Data Availability

All data produced in the present study are available upon reasonable request to the authors

## Acknowledgments

This study was funded by grants from NIH P2CHD086844 Medical University of South Carolina’s National Center of Neuromodulation for Neurorehabilitation and UPMC Rehabilitation Institute

